# Academic institution extensive, building-by-building wastewater-based surveillance platform for SARS-CoV-2 monitoring, clinical data correlation, and potential national proxy

**DOI:** 10.1101/2024.09.05.24313081

**Authors:** Arnoldo Armenta-Castro, Mariel Araceli Oyervides-Muñoz, Alberto Aguayo-Acosta, Sofia Liliana Lucero-Saucedo, Alejandro Robles-Zamora, Kassandra O. Rodriguez-Aguillón, Antonio Ovalle-Carcaño, Roberto Parra-Saldívar, Juan Eduardo Sosa-Hernández

**Author notes:** Corresponding authors, (R.P.-S.);, (J.E.S.-H).

## Abstract

Wastewater-based surveillance has been proposed as a cost-effective toolset to generate data about public health status by detecting specific biomarkers in wastewater samples, as shown during the COVID-19 pandemic. In this work, we report on the performance of an extensive, building-by-building wastewater surveillance platform deployed across 38 locations of the largest private university system in Mexico, spanning 19 of the 32 states. Sampling took place weekly from January 2021 and June 2022. Data from 343 sampling sites was clustered by campus and by state and evaluated through its correlation with the seven-day average of daily new COVID-19 cases in each cluster. Statistically significant linear correlations (p-values below 0.05) were found in 25 of the 38 campuses and 13 of the 19 states. Moreover, to evaluate the effectiveness of epidemiologic containment measures taken by the institution across 2021 and the potential of university campuses as representative sampling points for surveillance in future public health emergencies in the Monterrey Metropolitan Area, correlation between new COVID-19 cases and viral loads in weekly wastewater samples was found to be stronger in Dulces Nombres, the largest wastewater treatment plant in the city (Pearson coefficient: 0.6456, p-value: 6.36710^−8^), than in the largest university campus in the study (Pearson coefficient: 0.4860, p-value: 8.288×10^−5^). However, when comparing the data after urban mobility returned to pre-pandemic levels, correlation levels in both locations became comparable (0.894 for the university campus and 0.865 for Dulces Nombres).

## 1. Introduction

In recent years, wastewater-based surveillance (WBS) has emerged as a powerful toolset to provide data regarding public and environmental health status in communities through the detection and quantification of specific biomarkers in the sewage system (Choi et al., 2018, Lorenzo and Picó, 2019). While initial studies focused on chemical indicators of exposure to drugs, pharmaceuticals, contaminants, and personal care products, among others (Vitale et al., 2021), WBS has proven valuable for the study of the epidemiology of infectious diseases, such as Hepatitis E (Beyer et al., 2020), Norovirus (Guo et al., 2022), and other enteric viruses (Janahi et al., 2021). However, WBS rose to prominence during the COVID-19 pandemic, when it proved valuable for the prevention and contention of outbreaks by detecting and quantifying viral genetic material using RT-PCR-based methods (Bivins et al., 2020). It must be noted that WBS data has proven to be significantly more useful when used in tandem with clinical reports, leading to robust risk assessment models that can be used to predict surges in cases, allowing for well-informed decision-making (Islam et al., 2023).

In WBS platforms, sampling usually happens at wastewater treatment plants (WWTPs), since municipal wastewater can be used as a representative sample of its served population, allowing for reduced bias in the resulting datasets (Jimenez-Rodríguez et al., 2022). Nevertheless, several studies have explored the potential of targeted systems to study the circulation of pathogens within specific, high-affluence areas, such as schools, hospitals, college campuses (Gonçalves et al., 2022), and airports (van der Drift et al., 2024). As discussed by Wolken et al. (2023) strategic, targeted approaches are useful as a closer representation of the health status of subpopulations within a city and to prevent these buildings from becoming transmission hotspots, allowing for safer operations in the context of a pandemic. Studies where proved to be effective as an effective early warning system in the case of surges of COVID-19 cases, as sensitive and specific detection of viral genetic materials allowed for detection of both symptomatic and asymptomatic cases (Scott et al., 2021; Gibas et al., 2021; Corchis-Scott et al., 2021; Wang et al., 2022; Godinez et al., 2022). Moreover, longitudinal surveillance data can be used to reduce the intensity of individual clinical testing in periods when on-campus activities were returning to normalcy (Betancourt et al., 2021; Amirali et al., 2024), and to evaluate the efficacy of mitigation measures, such as social distancing, compulsive facemask use (Rainey et al., 2023), and vaccination (Bivins and Bibby, 2021), among others.

Previous efforts by our team explored the implementation of a WBS platform to survey the SARS-CoV-2 viral loads in the WWTPs of the Monterrey Metropolitan Area (MMA) in northern Mexico, correlating them with trends in clinical reports and urban mobility (Sosa-Hernández et al., 2022). In the present work, we implemented a similar platform to report the diversity and abundance of SARS-CoV-2 variants of concern in wastewater samples from the Mexico City’s sewage system (Aguayo-Acosta et al., 2024). We now report on the deployment and evaluation of a comprehensive WBS platform for the surveillance of SARS-CoV-2 on a building-by-building level across all the facilities of the largest private university in Mexico between January 2021 and June 2022. The institution operates 38 facilities in 19 of the 32 states of the country, serving 96,040 students as of 2022 (Tecnológico de Monterrey, 2022).

This study’s objectives were to establish an extensive, building-by-building WBS platform across all facilities and to evaluate the correlation between the load of viral genetic materials in wastewater samples with epidemiologic reports. Finally, to evaluate the effectiveness of the mitigation measures taken by the institution, results from the central university campus, located in the MMA (Campus Monterrey from now on) will be explored and compared to data originating from wastewater samples from Dulces Nombres, the largest WWTP operating within the MMA.

## 2. Methods

### 2.1. Wastewater sampling protocols

Wastewater sampling took place in 343 buildings across 39 participating facilities, including high schools, university campuses, and hospitals. The geographical distribution of campuses sampled in this study is shown in Figure 1. Samples were collected once a week between January 29, 2021 (denoted as week 5 from now on) and June 20, 2022 (denoted as week 78 from now on) in the wastewater discharge point of each participant building. Simple, 1 L grab samples were collected using high-density polyethylene (HDPE) bottles and stored at 4° C in ice packs for shipping to the central WBS laboratory, located within Campus Monterrey. In parallel, wastewater sampling was also conducted weekly at Dulces Nombres, the largest WWTP in the MMA, with a capacity of 7,500 liters per second and a served population of 1,695,589 inhabitants, in continuation of previous efforts by our team (Sosa-Hernández et al., 2022).

**FIGURE 1.**
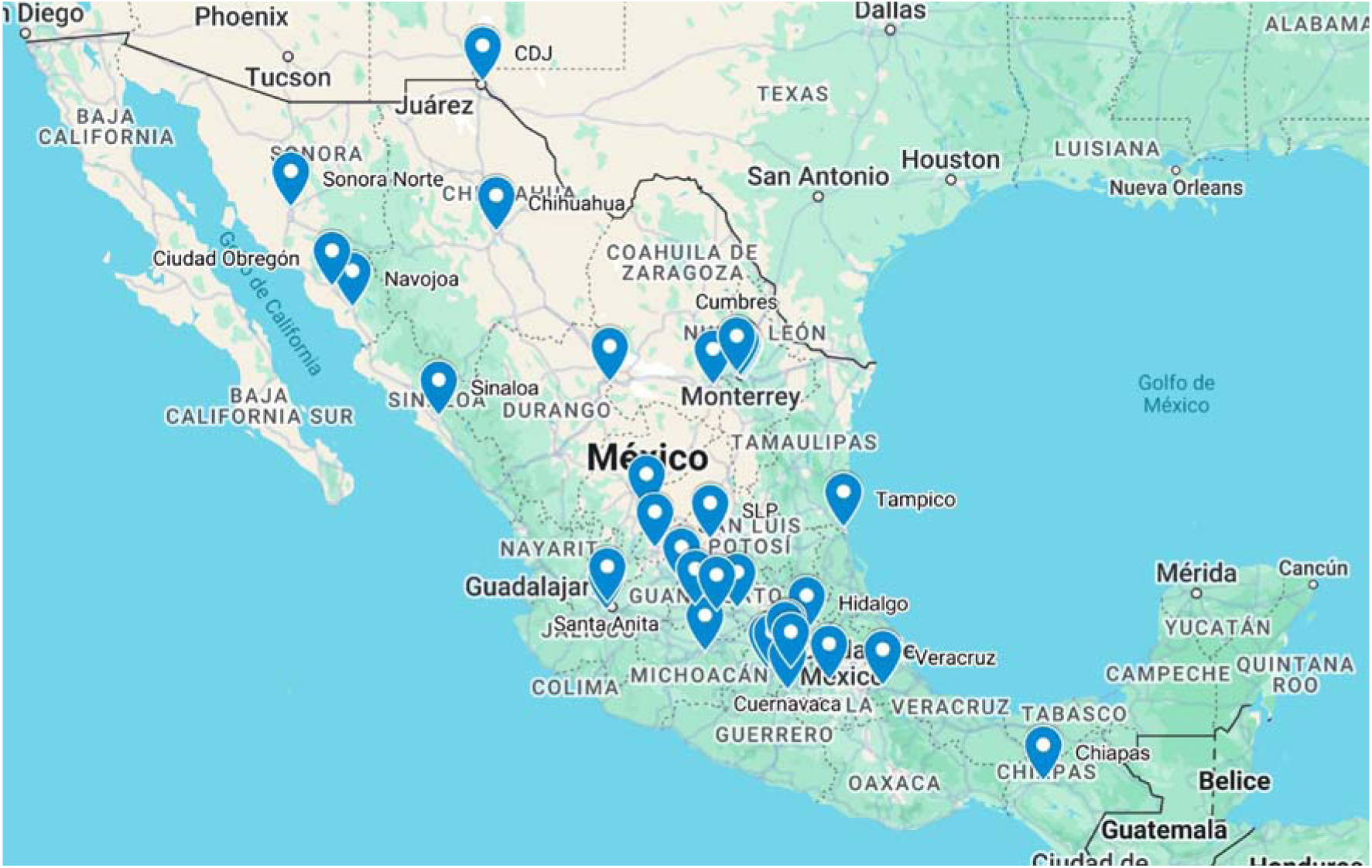
Geographical distribution of participating sampling sites across Mexico.

### 2.2. Concentration and extraction of genetic materials

Immediately after reception, samples were pre-processed to concentrate viral genetic materials utilizing a PEG/NaCl precipitation method based on the one reported by Sapula et al. (2021). Briefly, 70 mL from each sample was collected in two, 50 mL Falcon tubes and clarified by centrifugation at 5,000 g for 5 minutes. Supernatants were collected into new tubes with a polyethylene glycol 8000 and NaCl solution, shaken manually until the phases were homogenized, and the genetic materials were precipitated by centrifugation at 12,000 g for 100 minutes. After discarding the liquid phase, pelleted genetic materials were resuspended in 300 µL of Milli-Q water and stored at –20 °C until extraction.

Extraction was conducted using a Water DNA/RNA Magnetic Bead Kit (IDEXX, Westbrook, Maine) adapted for automation using a KingFisher™ Flex instrument (Thermo Fisher, Waltham, Massachusetts) following the supplier’s guidelines. Final elution volumes were kept at 100 µL in all cases. RT-PCR detection of SARS-CoV-2 genetic materials was conducted on a QuantStudio 5 instrument (Applied Biosystems, Waltham, Massachusetts) using the SARS-CoV-2 RTPCR Test kit for wastewater samples (IDEXX, Westbrook, Maine). In accordance with the supplier’s guidelines, reactions consisted of 5 µL of SARS-CoV-2 mix, 5 µL of RNA Master Mix, and 5 µL of extracted genetic material. The RT-PCR program consisted of an initial hold for 15 min at 50 °C and then 1 min at 95 °C, followed by 45 amplification cycles of 95 °C for 15 s and 60 °C for 30 s. Ct values were collected for each sample into a database for analysis.

### 2.3. Epidemiological clinical data acquisition

Total daily reported COVID-19 cases by the state were obtained from the dashboard published by the National Council of Humanities, Sciences, and Technologies (CONAHCYT) with data provided by the General Direction of Epidemiology, a part of the Mexican Department of Health (available at https://datos.covid-19.conacyt.mx/). As WBS data was collected weekly, a seven-day average of daily cases was calculated for each state across the period of study. Fluctuations in urban mobility in the MMA due to social distancing and other mitigation policies were accounted for using COVID-19 Community Mobility Reports published by Google in 2021 and 2022 for the Mexican state of Nuevo León (available at https://www.google.com/covid19/mobility/). Total changes in mobility were estimated by averaging the data from the six reported parameters (retail and recreation, groceries and pharmacies, parks, transit stations, workplaces, and residential). Finally, as this data was also reported daily, a seven-day average was paired to the weekly WBS data originating from Campus Monterrey for analysis.

### 2.4. Data analysis and visualization

WBS and epidemiological data was originally captured and organized in spreadsheets using Microsoft Excel, saved as CSV files, and imported into Matlab R2024a for visualization and statistical analysis. To limit the impact of inconsistencies during sampling due to restrictions at the sites or limited personnel, sites were clustered first by campus and by state for analysis. Four parameters were calculated weekly for each cluster: the average viral load of the weekly samples from each site (calculated as the sum of all the detected viral loads, expressed as copies per liter, divided by the number of samples taken from the site each week), the maximum viral load detected on the weekly samples from each site, the proportion of weekly positive samples (calculated as the number of positive samples from a site divided the total number of samples each week), and the proportion of buildings where the viral load was detected (calculated as the number of weekly positive samples divided by the total number of buildings sampled in the site).

Pearson correlation coefficients and their associated p-values were calculated for the seven-day average daily new COVID-19 cases in the corresponding state and each of the four parameters discussed above to evaluate the concordance of results obtained from the detection of SARS-CoV-2 genetic materials in wastewater samples with the evolution of the pandemic in the populations around each of the campuses studied. All correlations with a p-value below 0.05 were considered statistically significant.

For the state of Nuevo León, a comparative analysis between the data obtained from Campus Monterrey and Dulces Nombres was performed to evaluate the efficacy of the preventive measures taken by the institution. For this, Pearson correlation coefficients between the seven-day average new daily COVID-19 cases and the maximum viral load detected at Campus Monterrey and at Dulces Nombres, respectively, using both data from the entire study period and data obtained after urban mobility returned to pre-pandemic levels.

## 3. Results and discussion

### 3.1. Overview of wastewater sampling at Tecnológico de Monterrey

A total of 9664 wastewater samples were collected, processed, and analyzed in this study. However, as seen in Table 1, the distribution of sampling sites across facilities was not homogeneous due to differences in area, the number of buildings and the size of the student population in each one. The campuses from where most samples were collected were Monterrey (1395 samples), Querétaro (975), Guadalajara (634), Laguna (619), and León (551), all of them located in large metropolitan areas across Mexico. Similarly, the states from which more samples originated from were Nuevo León (2006 samples), Mexico (987), Querétaro (975), Chihuahua (879), and Mexico City (855). As observed previously in similar efforts, such as the one reported by Wolken et al. (2023), weekly sampling was not perfectly consistent due to factors including inaccessible sampling points, temporary closing of buildings due to surges in cases, lack of trained personnel, and scheduled holiday periods. As a result, while the total sampling period across the study extended for 74 weeks, the actual number of weeks in which sampling took place at Tecnológico de Monterrey facilities ranged from 62 at Campus Puebla to 14 at Hospital Zambrano. Likewise, the number of weeks in which sampling took place in at least one site in each state went from 62 in Puebla to 43 in Sinaloa. It must be noted that a total of two samples from Campus Veracruz were taken at the beginning of the study, but no further sampling was possible since the facility was closed shortly after. As such, it will not be considered in further data analysis.

**Table 1.** General summary of samples taken during the study, organized by campus and state of origin.

| No. | Site | Total samples | Weekly samplings | Buildings |
| --- | --- | --- | --- | --- |
| **Per campus** |  |  |  |  |
| 1 | Monterrey | 1395 | 60 | 42 |
| 2 | Querétaro | 975 | 60 | 25 |
| 3 | Guadalajara | 634 | 60 | 28 |
| 4 | Laguna | 619 | 56 | 15 |
| 5 | León | 551 | 61 | 13 |
| 6 | Chihuahua | 502 | 60 | 14 |
| 7 | Irapuato | 378 | 59 | 10 |
| 8 | CDJ | 377 | 60 | 14 |
| 9 | Estado de México | 373 | 55 | 18 |
| 10 | Zacatecas | 337 | 51 | 8 |
| 11 | Puebla | 336 | 62 | 11 |
| 12 | CDMX | 281 | 60 | 8 |
| 13 | Aguascalientes | 258 | 61 | 9 |
| 14 | Saltillo | 236 | 52 | 7 |
| 15 | Sonora Norte | 228 | 57 | 10 |
| 16 | Sinaloa | 199 | 43 | 9 |
| 17 | Toluca | 194 | 52 | 8 |
| 18 | SLP | 158 | 56 | 6 |
| 19 | Tampico | 152 | 58 | 7 |
| 20 | Santa Fe | 140 | 57 | 9 |
| 21 | Santa Catarina | 139 | 47 | 4 |
| 22 | Hidalgo | 134 | 59 | 6 |
| 23 | Cumbres | 133 | 52 | 4 |
| 24 | Morelia | 112 | 58 | 7 |
| 25 | Chiapas | 98 | 59 | 4 |
| 26 | Eugenio Garza Lagüera | 90 | 45 | 3 |
| 27 | Navojoa | 88 | 54 | 5 |
| 28 | Hospital Zambrano Hellion | 70 | 14 | 5 |
| 29 | Celaya | 58 | 58 | 4 |
| 30 | Ciudad Obregón | 58 | 54 | 4 |
| 31 | Cuernavaca | 57 | 53 | 5 |
| 32 | Esmeralda | 56 | 53 | 4 |
| 33 | Hospital San José | 51 | 17 | 4 |
| 34 | Valle Alto | 50 | 44 | 4 |
| 35 | Metepec | 43 | 43 | 2 |
| 36 | Eugenio Garza Sada | 42 | 42 | 3 |
| 37 | EGADE y EGOB | 36 | 36 | 1 |
| 38 | Santa Anita | 24 | 24 | 2 |
| 39 | Veracruz | 2 | 2 | 1 |

| Per state |  |  |  |  |
| --- | --- | --- | --- | --- |
| 1 | NUEVOLEON | 2006 | 61 | 59 |
| 2 | MEXICO | 987 | 61 | 23 |
| 3 | QUERETARO | 975 | 60 | 27 |
| 4 | CHIHUAHUA | 879 | 61 | 28 |
| 5 | DISTRITOFEDERAL | 855 | 57 | 21 |
| 6 | HIDALGO | 658 | 60 | 27 |
| 7 | JALISCO | 610 | 55 | 26 |
| 8 | COAHUILA | 477 | 61 | 20 |
| 9 | SONORA | 374 | 57 | 15 |
| 10 | ZACATECAS | 337 | 51 | 8 |
| 11 | PUEBLA | 336 | 62 | 11 |
| 12 | AGUASCALIENTES | 258 | 61 | 9 |
| 13 | SINALOA | 199 | 43 | 9 |
| 14 | SANLUISPOTOSI | 158 | 56 | 6 |
| 15 | TAMAULIPAS | 152 | 58 | 10 |
| 16 | GUANAJUATO | 134 | 59 | 6 |
| 17 | MICHOACAN | 112 | 58 | 7 |
| 18 | CHIAPAS | 98 | 59 | 5 |
| 19 | MORELOS | 57 | 53 | 5 |
| 20 | VERACRUZ | 2 | 2 | 1 |

Similarly, a summary of the sampling results is presented in Figure 2A for campuses and Figure 2B for states. Red squares show weeks when sampling was conducted at a given site and at least one of the sampled buildings tested positive for SARS-CoV-2 genetic materials, white squares show weeks when sampling was conducted but no viral loads were detected, and grey squares show weeks when no sampling was conducted As a reference, a graph of the weekly evolution of new COVID-19 cases in Mexico is reported in Figure 2C. Complete records of all samples collected during the study, their origin, and the detected viral loads, reported as copies per liter of the original wastewater sample, are available in the Supplementary Materials.

**Figure 2.**
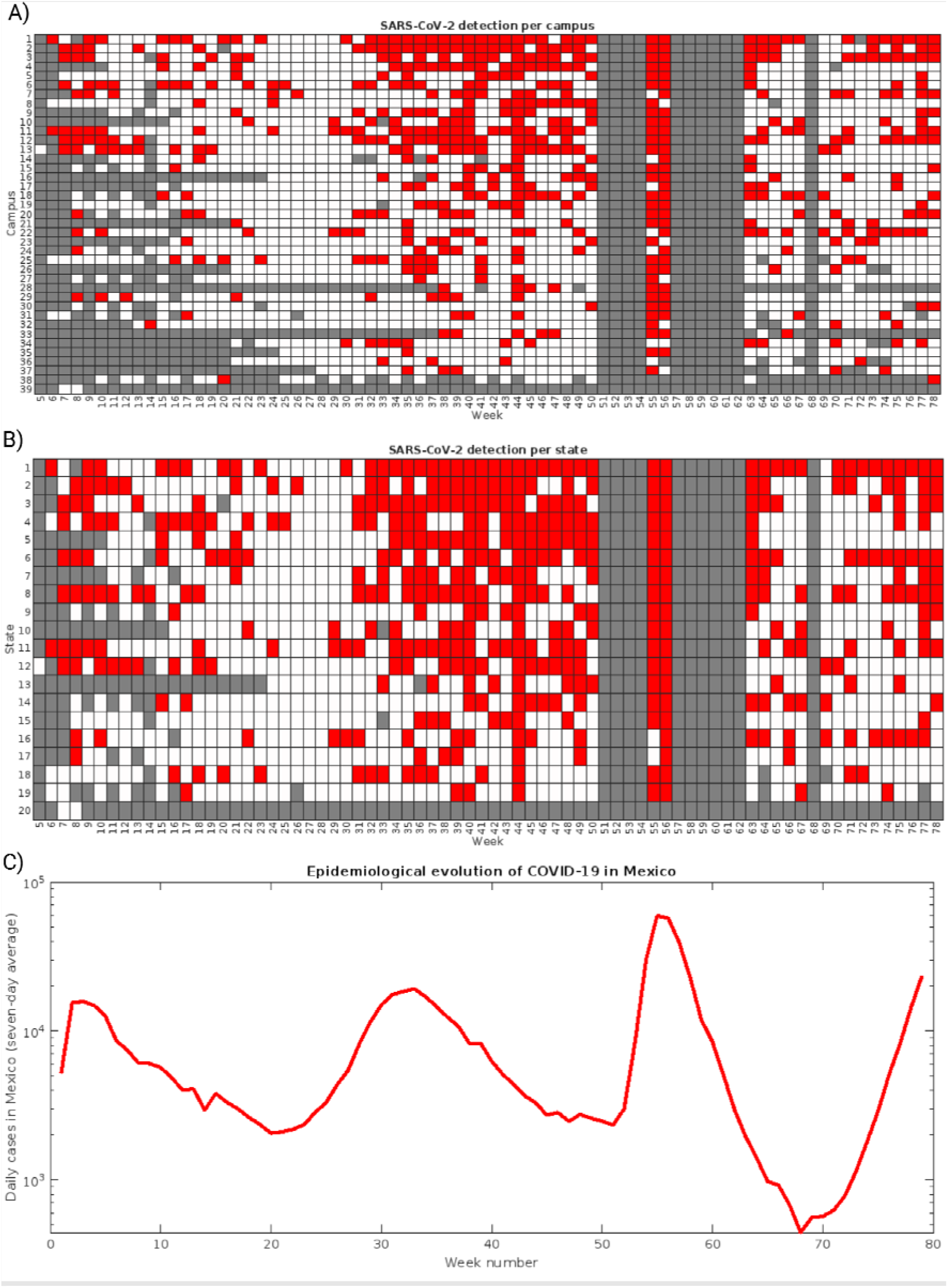
Summary of samplings across participant sites. A) By campus, B) by state. C) Weekly evolution of new daily COVID-19 cases across Mexico.

### 3.2. Correlation between WBS results and epidemiological reports

As shown in Table 2, the maximum viral load and the average viral load in the weekly samples taken in each site showed the most consistent correlation to the seven-day average of new daily COVID-19 cases in the corresponding state (available in the Supplementary Materials), both when campuses were analyzed individually (average correlation coefficients of 0.448 ± 0.302 and 0.427 ± 0.303, respectively) and when campuses were clustered by state (0.465 ± 0.276 and 0.438 ± 0.277). The correlation with the proportion of positive samples and positive buildings fell significantly (0.296 ± 0.197 and 0.309 ± 0.201 for individual campuses, 0.307 ± 0.153 and 0.317 ± 0.180 for states), likely due to the inconsistencies in samples discussed previously, which led to significant variations in the number of samples being obtained each week. The number of sites where correlation proved statistically significant (p-values below 0.05) showed similar behavior: correlation to the maximum viral load per site showed to be significant in 25 of the 38 campuses and 13 of the 19 states, followed by the average viral load per campus or state (24/38 and 13/19, respectively).

**Table 2.** Average correlation coefficients for the average viral load, maximum viral load, proportion of positive samples, and proportion of positive buildings across sampling sites, by campus and by state.

|  | Correlation with seven-day average new daily cases |  |  |  |
| --- | --- | --- | --- | --- |
|  | Average viral load | Maximum viral load | Proportion of positive samples | Proportion of positive buildings |
| Per campus |  |  |  |  |
| Average correlation coefficient (Pearson) | $0.427 \pm 0.303$ | $0.448 \pm 0.302$ | $0.296 \pm 0.197$ | $0.309 \pm 0.201$ |
| Sites with statistically significant correlation | 24 | 25 | 22 | 22 |
| Percentage of sites with statistically significant correlation | 63.16% | 65.79% | 57.89% | 57.89% |

| Per state |  |  |  |  |
| --- | --- | --- | --- | --- |
| Average correlation coefficient (Pearson) | 0.438 ± 0.277 | 0.465 ± 0.276 | 0.307 ± 0.153 | 0.317 ± 0.180 |
| Sites with statistically significant correlation | 13 | 13 | 12 | 11 |
| Percentage of sites with statistically significant correlation | 68.42% | 68.42% | 63.16% | 57.89% |

As suggested previously, correlation coefficients vary greatly among campuses, going from 0.9004 at Hospital San José to −0.0942 in Campus Sinaloa, although most campuses show positive correlation coefficients and 24 out of 38 show coefficients above 0.4. The complete list is presented in Table 3 of number of weekly samplings and the average coverage rate (calculated as the average of the ratios between samples taken on each campus per week and the total number of sampling sites within the campus) for each campus, the Pearson correlation between the maximum viral load found weekly on each campus and the seven-day average of new daily COVID-19 cases in the state where the campus is located, and its related p-value.

**Table 3.**
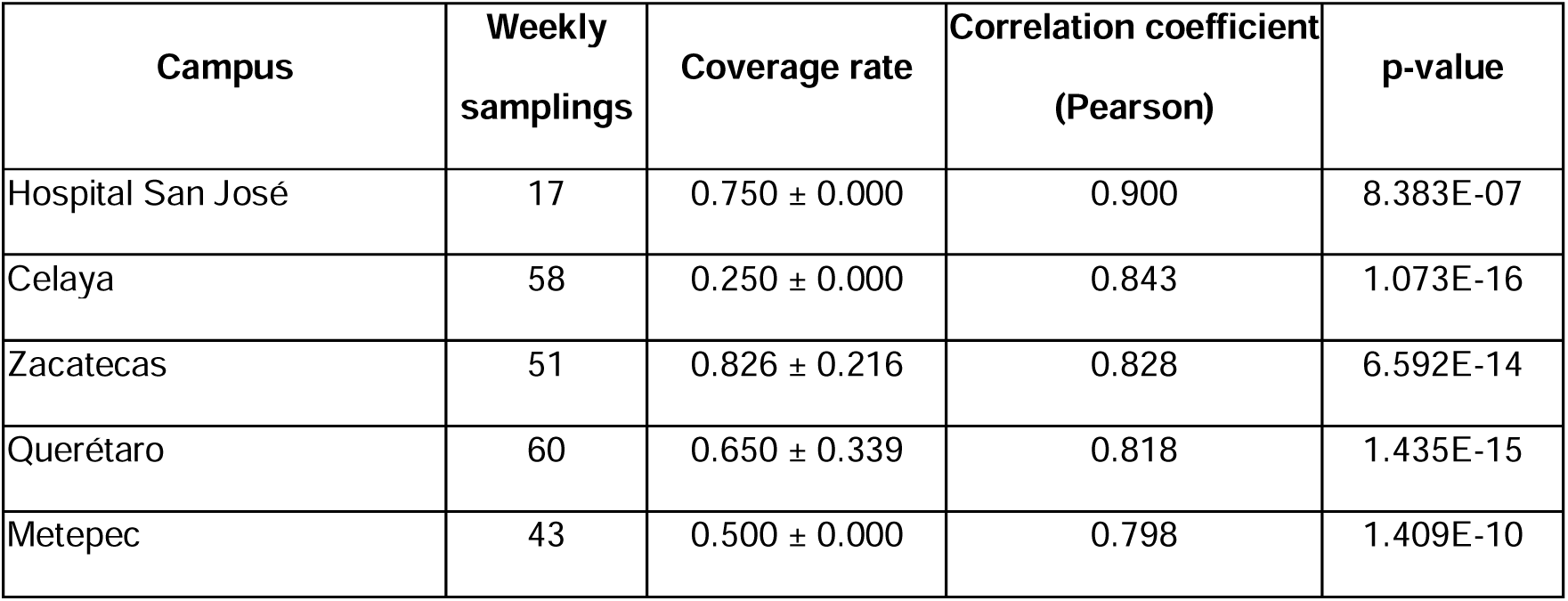
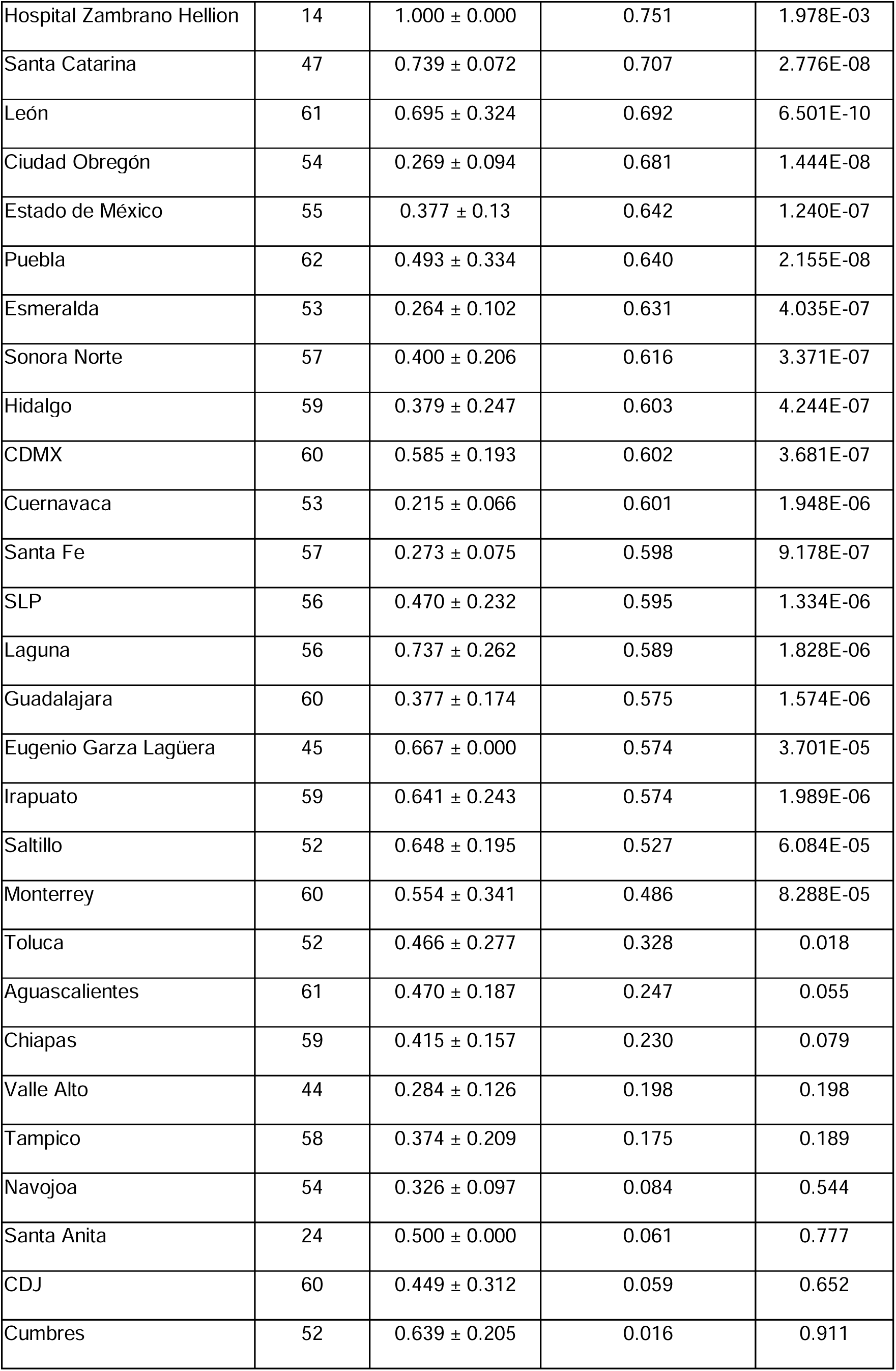
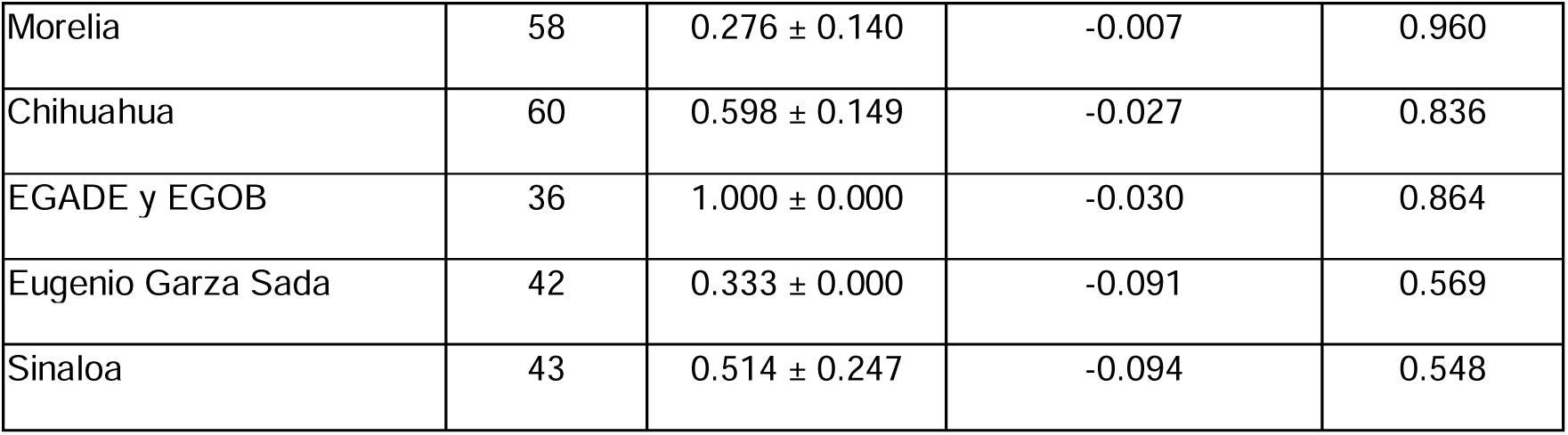
Correlation coefficients between the maximum viral load and the seven-day average of new daily COVID-19 cases across campuses.

Just as observed when clustering sampling sites by campus, correlation coefficients vary greatly among states, going from 0.8280 in Zacatecas to −0.0942 in Sinaloa, although most states show positive correlation coefficients, and 13 out of 19 show coefficients above 0.4. The complete list is presented in Table 4 with the number of weekly samplings and the average coverage rate (calculated as the average of the ratios between samples taken on each state per week and the total number of sampling sites within the campus) for each state, the Pearson correlation between the maximum viral load found weekly on each state and its seven-day average of new daily COVID-19 cases, and the related p-value.

**Table 4.** Correlation coefficients between the maximum viral load and the seven-day average of new daily COVID-19 cases across states.

| Campus | Weekly<br>sampling | Coverage rate | Correlation coefficient<br>(Pearson) | p-value |
| --- | --- | --- | --- | --- |
| ZACATECAS | 51 | 0.826 ± 0.216 | 0.828 | 6.592E-14 |
| QUERETARO | 60 | 0.650 ± 0.339 | 0.818 | 1.435E-15 |
| GUANAJUATO | 61 | 0.952 ± 0.398 | 0.657 | 8.834E-09 |
| MEXICO | 55 | 0.616 ± 0.255 | 0.643 | 1.208E-07 |
| PUEBLA | 62 | 0.493 ± 0.334 | 0.640 | 2.155E-08 |
| SONORA | 57 | 0.596 ± 0.232 | 0.635 | 1.143E-07 |
| COAHUILA | 61 | 0.711 ± 0.239 | 0.632 | 4.829E-08 |
| HIDALGO | 59 | 0.379 ± 0.247 | 0.603 | 4.244E-07 |
| MORELOS | 53 | 0.215 ± 0.066 | 0.601 | 1.948E-06 |
| SAN LUIS POTOSI | 56 | $0.470 \pm 0.232$ | 0.595 | 1.334E-06 |
| DISTRITO FEDERAL | 57 | $0.882 \pm 0.306$ | 0.589 | 1.445E-06 |
| JALISCO | 60 | $0.392 \pm 0.171$ | 0.575 | 1.575E-06 |
| NUEVO LEON | 61 | $0.747 \pm 0.451$ | 0.486 | 7.067E-05 |
| AGUASCALIENTES | 61 | $0.47 \pm 0.187$ | 0.247 | 0.055 |
| CHIAPAS | 59 | $0.415 \pm 0.157$ | 0.230 | 0.079 |
| TAMAULIPAS | 58 | $0.374 \pm 0.209$ | 0.175 | 0.189 |
| MICHOACAN | 58 | $0.276 \pm 0.14$ | -0.007 | 0.960 |
| CHIHUAHUA | 61 | $1.029 \pm 0.432$ | -0.018 | 0.889 |
| SINALOA | 43 | $0.514 \pm 0.247$ | -0.094 | 0.548 |

Limited correlation between the results of the WBS platform deployed across participating facilities and the epidemiological reports was expected, as measures to prevent SARS-CoV-2 transmission, including mandatory face mask-wearing, vaccination, and reduced levels of in-person activities were implemented across the entire period of study, altering the population dynamics, viral transmission dynamics, transmission risk and density of exposure in campus, as studied by Tsang et al. (2023), affecting the correlation observed between viral load in campus wastewater and daily cases of COVID-19 in MMA. In fact, WBS reports obtained by our research team were used by officials to limit the activities in buildings that proved positive for SARS-CoV-2 genetic materials in a mechanism like the one reported by Wang et al. (2022). However, estimating the efficacy of outbreak prevention measures using WBS data has proven difficult, as adequately modeling the rates of viral shedding (Zhu et al., 2021) and RNA degradation in sewage systems (Parra-Arroyo et al., 2023) has been identified as a challenge. Moreover, reports on the implementation of extensive, targeted surveillance platforms, such as the ones by Bowes et al. (2023) and Wolken et al. (2023), have stressed the importance of adequate standardization of sampling, pretreatment, concentration and extraction procedures, as well as the proper identification the sensitivity, specificity and limit of detection of the technique used for SARS-CoV-2 viral load detection to minimize systematic variation and obtain complete, robust data sets.

While the performance of the building-by-building WBS platform was satisfactory, it is important to highlight the challenges related to the operation of such a large network of sampling sites distributed in a vast geographical area, including incomplete sample collection due to unforeseeable circumstances during the study period and inadequate sampling handling during transportation, likely a result of insufficient cold chains, as reviewed by Bengiovanni et al. (2020). These obstacles led to the observed variation in the number of weekly samplings taken per campus and by state, in the effective coverage rates, and the representativity obtained in each campus when compared to the epidemiological data reported by the public health authorities. Extensive networks would be ideal to obtain more detailed information that could be used for better risk assessment models, especially in areas that are particularly prone to disease outbreaks (Gonçalves et al., 2022); however, limited datasets are to be expected as surveillance platforms expand, especially in Low-to-Middle Income Countries like Mexico, where such systems are often operated with limited resources (Shrestha et al., 2021; Hamilton et al., 2024). Similar surveillance efforts conducted in the future would likely benefit from a more targeted approach, where sampling sites are set strategically, to allow for more sustained, consistent coverage while retaining sample representativity. In cases of public health emergencies where extensive, sustained surveillance is crucial for public health, as in the case of the COVID-19 pandemic, it is likely that WBS platforms would be more effective when operated collaboratively across institutions as long as the procedures for sample handling, pretreatment, and viral genetic material extraction and detection are properly standardized and proven reproducible.

### 3.3. Comparisons between data from a college campus and a WWTP

To offer insights on the effectiveness of the epidemiological contention and prevention taken by Tecnológico de Monterrey (including reduced in-person activities, compulsive mask wearing and vaccination), data obtained from wastewater surveillance in Campus Monterrey and the Dulces Nombres WWTP (avaible in the Supplementary Materials), both located in the MMA, was compared to epidemiological data reported by public health authorities for the state of Nuevo León. After eliminating weeks when no sampling took place, correlation between the maximum viral load found on all the samples originating from Campus Monterrey each week and the corresponding seven-day average of new daily cases of COVID-19 reported in Nuevo León was 0.486 (p-value: 8.2876×10^−5^). For the viral load detected at Dulces Nombres, the observed correlation coefficient reached 0.6356 (p-value: 6.3672^−8^). Plots presenting the distribution of the data from each weekly sampling is presented in Figure 3A.

**Figure 3.**
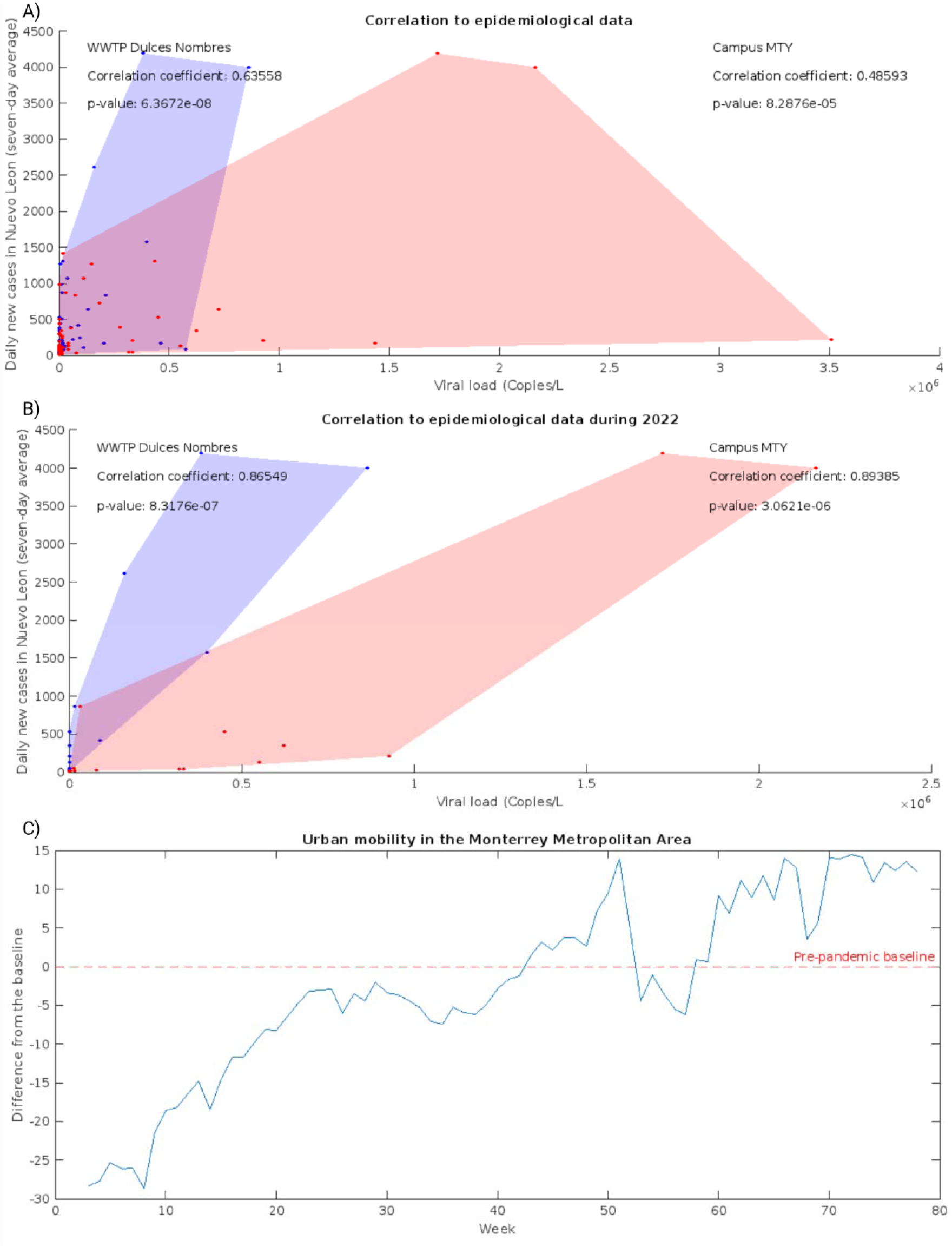
Correlation between the seven-day average of daily new cases of COVID-19 in the state of Nuevo León and the viral load detected at Campus Monterrey (blue) and Dulces Nombres (red). A) Including the entire period of study. B) Only considering data from January to June 2022. C) Evolution of urban mobility in the MMA across the period of study.

Since samples from both sites originate from the same metropolitan area and were transported a similar distance to the central laboratory following standard practices for genetic material conservation, it can be suggested that the difference in their degree of correlation to the epidemiological situation in the state is due to the different populational dynamics they represent. Campus Monterrey, like most higher education facilities in Mexico, greatly limited its in-person activities during the pandemic; maintenance personnel, researchers, postgraduate students, and security guards were still going regularly into the facilities, but most of the undergraduate students, faculty, and administrative personnel conducted their activities remotely. Inversely, Dulces Nombres is the biggest collection point for domestic and industrial wastewater in the MMA and, as a result, is representative of the entire population within its catchment area, regardless of whether they adhered to social distancing regulations, presented any symptoms of COVID-19, or received a clinical test and were reported to the epidemiological databased compiled and published by public health authorities. Therefore, it can be observed that, while extensive wastewater surveillance at Campus Monterrey was useful for a safe continuation of strategic activities at campus that could not be conducted remotely, WBS platforms centered on WWTPs were still more representative of the epidemiological situation in the MMA, where the implementation of transmission measure prevention doesn’t impact in wastewater viral load, contrary to what has been observed on the campuses.

In a related line, Figure 3B shows the evolution of urban activity in the state of Nuevo Leon during the period of study as reported in the COVID-19 Community Mobility Reports published by Google. Weeks are presented in the X-axis, while changes in mobility compared to the baseline (from January 3 to February 6, 2020) are presented in the Y-axis. Mobility surpassed the pre-pandemic baseline in week 43 (the last week of October 2021) and remained above the baseline throughout 2022 (week 52 onwards) with the exceptions of weeks 53-57 (in January 2022), likely due to the holiday season and increased cases in the winter. This indicates a relative return to normalcy in the populational dynamics of the state. Taking this into account, correlations between the maximum viral load found each week at Campus Monterrey and at Dulces Nombres with the seven-day average of new daily cases in the state of Nuevo Leon between January and June of 2022. As expected, correlation increased noticeably in both cases due to less stringent epidemiological containment measures, reaching correlation coefficients of 0.8655 (p-value: 8.3176×10^−7^) for Dulces Nombres, and 0.8938 (p-value: 3.0621×10^−6^) for Campus Monterrey. Moreover, the fact that correlation coefficients for both sites were so close after week 52 indicates that the populational dynamics at Campus Monterrey became similar to MMA and more representative to the ones shown in the MMA and the state of Nuevo León, further suggesting the effectivity of epidemiological prevention taken at Campus Monterrey throughout 2021, which were implemented in conjunction with the rest of the facilities of Tecnológico de Monterrey across Mexico.

## 4. Perspectives

This work offers important insights into the operation of a large WBS platform for the surveillance of SARS-CoV-2 across 39 facilities across Tecnológico de Monterrey, encompassing high schools, college campuses, and hospitals in 20 of the 32 federal entities of Mexico. The performance of such a platform was evaluated through comparisons to epidemiological data reported by public health authorities and, in the case of Campus Monterrey (the largest of all participant facilities), to data obtained from samples taken at Dulces Nombres, the largest WWTP in the MMA. While it was observed that viral loads detected in wastewater samples were correlated with the amount of daily new COVID-19 cases in 25 campuses across 13 states, and that WBS could be used as an effective strategy to support epidemiological contention measures, as shown for the case of Campus Monterrey, opportunities for improvement of WBS platforms persist.

Sampling across a large network of sites across participating facilities was relevant for the institution, as it offered data to guide effective preventive efforts for a safe continuation of priority in-person activities. However, encompassing such a large set of sampling points into a single platform operated from a single laboratory made consistent coverage across campuses (defined here as the rate of total weekly samples from a facility over the total amount of participant buildings in the facility) difficult. While taking only the weekly maximum viral load for each campus for correlation studies helped with consistency when handling such an incomplete database, as shown by the performance metrics reported in the previous section, significant variance in coverage rates may have contributed to limited correlation between WBS data and epidemiological reports. Future efforts towards sustained, extensive surveillance platforms, either for detection of endemic pathogens such as Influenza or Norovirus, or future epidemiological outbreaks akin to SARS-CoV-2, may benefit from a more strategic selection of sampling points that can be studied consistently and yield results that can be representative of general populational dynamics, as previously suggested by Safford et al. (2022).

Moreover, standardized, reproducible sample processing methods will be crucial for any WBS platform, as inconsistent viral load detection and quantification may hinder data comparability and analysis, as discussed previously by Bowes et al. (2023). Moreover, levels of biomarkers in wastewater have been observed to vary significantly because of changes in the flow at the designated sampling site, fluctuations in human activity, and possible degradation of the molecule of interest (Ali et al., 2021). While the surveillance effort reported here relied on simple grab samples, a limitation that could be mitigated by clustering different sites by campus and by state during data analysis, smaller, more targeted platforms could benefit from the use of composite sampling or implementation of passive sampling technologies, although their applicability for wastewater surveillance of pathogens still requires further studies (Aguayo-Acosta et al., 2023).

Finally, integration of viral load quantification in wastewater samples into complete epidemiological models that could be used for risk assessment and prevention during a public health emergency, such as the one posed by SARS-CoV-2, remains as an area of opportunity for WBS. While certain directions for the interpretation of WBS data as an indicator of public health status have been discussed elsewhere (Islam et al., 2023), data normalization remains a challenge as no universal wastewater biomarker for population size and levels of human activity have been agreed upon, although several options have been suggested (Hsu et al., 2022). In any case, WBS should not be thought of as a substitute for individual clinical tests, but as a complement for more robust epidemiological data. This work focuses only on the main outcomes of the WBS platform deployed at Tecnológico de Monterrey, while their integration into epidemiological models will be explored in future studies.

## 5. Conclusions

Overall, the extensive, building-by-building WBS platform deployed across Tecnológico de Monterrey successfully reflected the evolution of the COVID-19 epidemic in 25 of the 38 facilities in the study and provided valuable insights for effective epidemiological containment during 2021, allowing for the continuation of priority in-person activities. Moreover, implementing a WBS platform targeting educative institutions like university campuses proved a suitable proxy to study the epidemiological dynamics in tandem with clinical reports published by public health authorities in larger communities, like states.

Additionally, this study evaluates the optimal strategy to obtain correlations between viral loads in wastewater samples from individual buildings and published clinical reports, proving that clustering sampling points by campus or state and using the maximum viral load found in each cluster is a feasible strategy to generate robust, insightful datasets for informed decision-making. In fact, data from Campus Monterrey became more representative of the public health condition in the state of Nuevo León after urban mobility went back to pre-pandemic levels at the start of 2022, indicating that the altered populational dynamics within the campus during 2021 due to limited in-person activities were effective to prevent COVID-19 outbreaks within the facility.

However, further studies regarding the optimization of such a vast platform and the integration of WBS data into broader epidemiological models are still needed for the development of robust, efficient surveillance platforms for future public health emergencies.

## Data Availability

The document has the data used in the SI section.

## Credit author statement

**Arnoldo Armenta-Castro:** Conceptualization, Formal analysis, Visualization, Methodology, Data Curation, Writing-Original draft preparation. **Alberto Aguayo-Acosta:** Conceptualization, Investigation, Formal analysis, Validation, Methodology, Writing-Original draft preparation, Writing - Review & Editing. **Mariel A. Oyervides-Muñoz:** Conceptualization, Investigation, Formal analysis, Validation, Methodology, Writing-Original draft preparation, Supervision, Writing - Review & Editing. **Sofia Liliana Lucero-Saucedo:** Investigation, Writing-Original draft preparation, Writing - Review & Editing. **Alejandro Robles-Zamora:** Investigation, Writing-Original draft preparation, Writing - Review & Editing. **Kassandra O. Rodriguez-Aguillón:** Investigation, Writing-Original draft preparation, Writing - Review & Editing. **Antonio Ovalle-Carcaño:** Investigation, Writing-Original draft preparation, Writing - Review & Editing. **Roberto Parra-Saldívar:** Conceptualization, Investigation, Writing-Original draft preparation, Supervision, Funding acquisition, Writing - Review & Editing. **Juan Eduardo Sosa-Hernández:** Conceptualization, Investigation, Writing-Original draft preparation, Supervision, Project administration, Funding acquisition, Writing - Review & Editing.

## Acknowledgments

The authors would like to thank the Bioproduction Systems and MARTEC lab from Tecnologico de Monterrey, Mexico. The authors appreciate the support of Tecnologico de Monterrey for granting access to literature services and the scholarship awarded to Arnoldo Armenta-Castro (CVU: 1275527) and partially supporting this work under Sistema Nacional de Investigadores program awarded to Alberto Aguayo-Acosta (CVU: 403948), Mariel A. Oyervides-Muñoz (CVU: 422778), Roberto Parra-Saldívar (CVU: 35753) and Juan Eduardo Sosa-Hernández (CVU: 375202).

